# Prioritising determinants of systemic inflammation across molecular, physiological and disease phenotypes

**DOI:** 10.64898/2026.04.10.26350510

**Authors:** Freya R. Shepherd, Chloe Slaney, Hannah Jones, Christina Dardani, Evie Stergiakouli, Eleanor C. M. Sanderson, Fergus Hamilton, Daniel B. Rosoff, Nils Rek, Tom R. Gaunt, George Davey Smith, Tom G. Richardson, Golam M. Khandaker

## Abstract

**Background:** Systemic inflammation has been widely studied as a driver of disease, yet its upstream determinants remain poorly understood.

**Methods:** We performed a large-scale two-sample Mendelian randomisation study evaluating potential causal effects of 3,213 molecular, physiological and disease traits on circulating interleukin-6 and C-reactive protein. These comprised 2,923 proteomic, 249 metabolomic, 17 cardiometabolic, 7 neuropsychiatric, 8 immune-mediated inflammatory and 9 infection-related traits. Genetic instruments from genome-wide association studies were analysed using inverse variance weighted, weighted median and MR-Egger methods, with multiple-testing correction. Consistency across methods informed robustness, and bidirectional MR assessed reverse causation.

**Results:** After Bonferroni correction, 72 traits showed evidence of potential causal effects on CRP and 9 on IL-6. CRP was predominantly influenced by metabolomic traits, particularly lipid and fatty acid measures. Genetically proxied adiposity (body mass index and obesity), triglyceride-rich lipoproteins, glycoprotein acetyls (GlycA) and apolipoprotein E increased CRP, whereas HDL-related cholesterols, polyunsaturated fatty acids and glutamine decreased CRP. Most associations were consistent across MR methods.

IL-6 was primarily influenced by adiposity and HDL-related lipid measures, with generally smaller effects and limited support from sensitivity analyses. Bidirectional analyses provided little evidence that CRP directly drives metabolic traits when restricted to cis-acting instruments, whereas genetically proxied IL-6 signalling showed consistent downstream effects on HDL particle concentration and composition.

**Conclusions:** Adiposity is a shared upstream determinant of both biomarkers, with stronger and broader effects on CRP. These findings suggest that CRP integrates downstream systemic inflammatory burden, whereas IL-6 reflects a more tightly regulated, context-dependent process. By shifting the focus from inflammation as a cause to inflammation as a consequence, this study maps potential causal drivers of systemic inflammation across a broad spectrum of traits and pathways, identifies pathways connecting inflammation with cardiometabolic and inflammatory diseases, and provides a resource for prioritising upstream determinants for mechanistic investigation and therapeutic targeting.

**Plain language summary:** Inflammation plays a vital role in protecting the body from infection and injury but can also become chronic and consequently detrimental. Systemic inflammation is linked to many common diseases, including heart disease, diabetes, and depression. C-reactive protein and Interleukin-6 are widely used markers of inflammation which can be measured in the blood. Although these markers are often elevated in disease, it is not always clear whether they are a cause or consequence of the disease, or related other underlying processes.

In this study, we used genetic evidence from large population studies to help clarify which traits may have causal influence on levels of these inflammatory markers. By analysing thousands of potential relationships across metabolic, immune, cardiovascular, and mental health traits, we identified several metabolic processes, and related traits such as BMI and Type 2 diabetes, as key drivers of both markers of inflammation. We also found that C-reactive protein appears to reflect a broader range of biological influences than Interleukin-6.

Our findings help to clarify which factors are most likely to sit upstream of systemic inflammation. This improved understanding may help guide future research aimed at preventing or reducing inflammation-related disease.

## 1. Introduction

An ever-increasing body of research implicates systemic inflammation in a wide variety of health conditions. Beyond infection [1] and immune mediated inflammatory disease [2], inflammation is now implicated in cardiometabolic [3] and neuropsychiatric conditions [4] and linked to changes in metabolomic [5] and proteomic parameters [6]. Interleukin-6 (IL-6) [7] and C-reactive protein (CRP) [1] are archetypal inflammatory markers and are two of the most extensively studied blood biomarkers in the context of health and disease.

IL-6 is a pleiotropic cytokine produced by immune and non-immune cells in response to infection, injury, and stress signals [8]. It regulates both pro- and anti-inflammatory pathways, influences immune cell differentiation, metabolism and acute phase responses [8]. Dysregulation of IL-6 signalling has been implicated in autoimmune disorders [8], cardiovascular disease [3] and depression [9]. CRP is an acute phase reactant synthesised by the liver, stimulated by IL-6 receptor (*IL-CR*) signalling [10]. Circulating CRP levels rise rapidly during inflammation and are widely used as a clinical biomarker of systemic inflammatory activity [10]. Elevated CRP concentrations have been consistently associated with increased risk of cardiovascular disease [11], metabolic syndrome [12], all-cause mortality [13], and several psychiatric conditions, notably depression [14] and schizophrenia [15].

Existing studies have largely focused on the role of CRP and IL-6 in relation to disease outcomes, but systematic examinations of molecular, physiological and disease phenotypes that causally drive circulating levels of these biomarkers are scarce. Mendelian randomisation (MR) is a genetic epidemiological approach that can assess evidence for causality [16,17] by using single nucleotide polymorphisms (SNPs) that are robustly associated with an exposure as proxies for that exposure [16]. SNPs are identified through genome wide association studies (GWAS) and are commonly used as proxies for an exposure of interest [18]; referred to instrumental variables (IVs).

To implement MR in an IV framework, several key assumptions must be met; the IVs must be robustly associated with the exposure, there can be no confounding of the IVs and the outcome, and the IVs must be independent of the outcome given the exposure and not via alternative biological pathways (the exclusion restriction assumption, i.e. absence of horizontal pleiotropy) [19]. Although MR is now commonly implemented within an instrumental variable (IV) framework using genetic variants, the concept of MR as a natural experiment predates its formalisation within the IV framework. In practice, MR inference relies on assessing the plausibility of core assumptions and the robustness of estimates to potential violations, rather than requiring these assumptions to be perfectly satisfied [20]. When the exposure is a binary or disease trait, MR estimates reflect the effect of genetic liability to that condition -modelled as an underlying continuous trait - rather than the effect of the diagnosed disease state itself on downstream outcomes [21].

By using the random allocation of alleles at conception and the fact that germline genetic variants are fixed across the life course, MR can reduce bias from confounding and reverse causation that commonly limit conventional observational studies, enabling causal inference between phenotypes using population-level data [16].

The primary objective of this study was to investigate whether genetically predicted molecular, physiological and disease-related traits have causal effects on circulating IL-6 and CRP concentrations. Exposures were grouped into domains selected based on prior evidence linking them to inflammation or immune dysfunction, including infectious and immune mediated inflammatory diseases, cardiometabolic and physiological traits, and molecular phenotypes derived from plasma proteomic and metabolomic data. By evaluating diverse molecular, metabolic, physiological, and disease-related traits, our analysis provides a systematic overview of potential upstream determinants of circulating IL-6 and CRP levels and highlights biological processes that may contribute to systemic inflammatory activation. This approach enables targeted systematic identification of upstream determinants of systemic inflammation, providing insight into shared biological pathways and potential pleiotropic influences.

To further assess causal directionality, we conducted targeted bidirectional MR analyses for traits showing significant associations in the primary MR analysis, testing whether IL-6 or CRP may in turn exert causal effects on these outcomes.

## 2. Methods

An overview of the analytical pipeline of the study can be found in **Figure 1**. Extended methods can be found at [22]. All GWAS summary statistics analysed in this study were sourced from previously published studies that had obtained ethical approval and informed consent from participants. A preprint version of this article is available [23].

**Figure 1.**
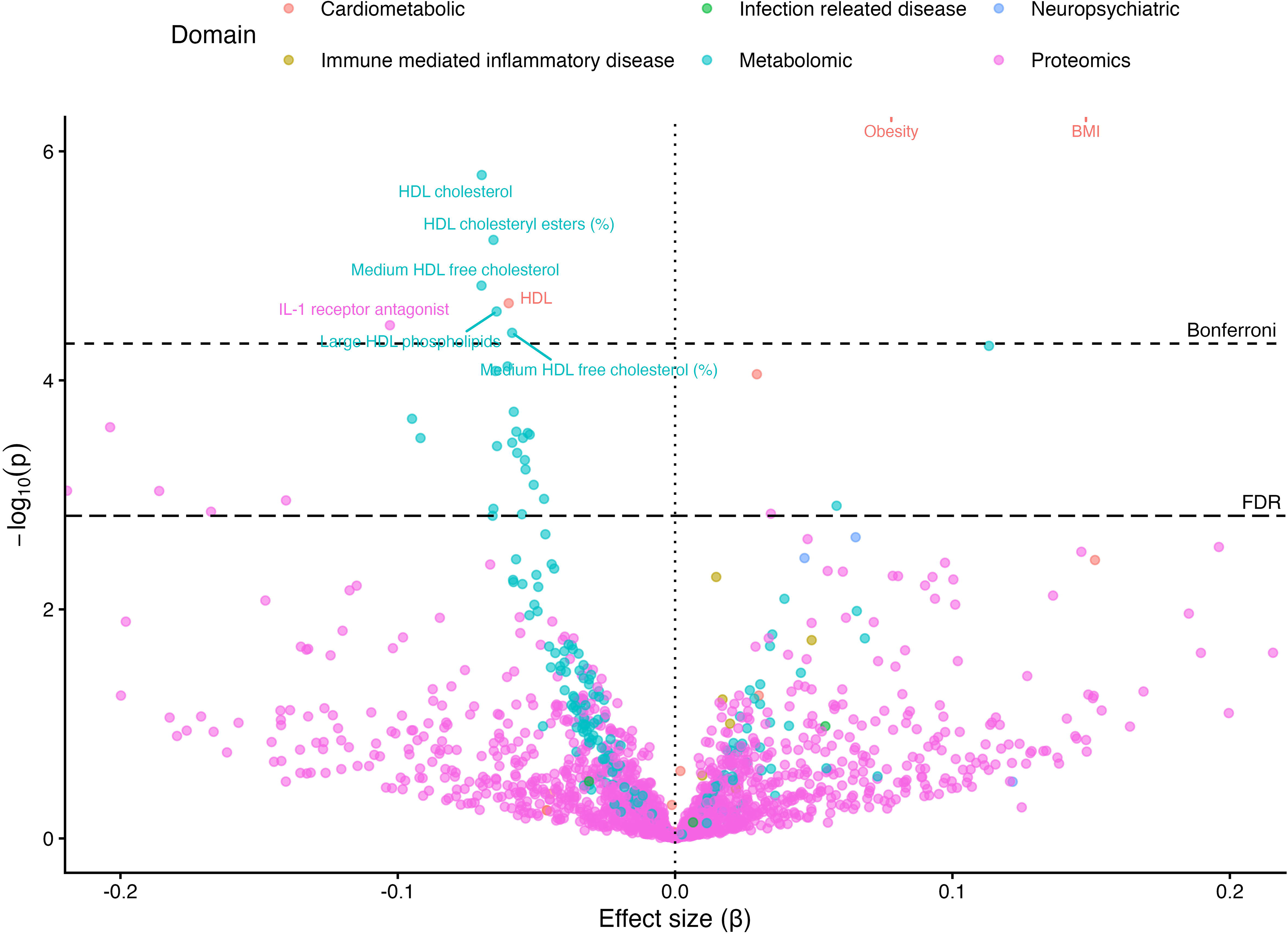
Overview of the analytical pipeline. Two-sample Mendelian randomisation (MR) was used to investigate potential causal effects of 3,213 targeted molecular, physiological and disease traits (exposures), grouped into six domains, on circulating interleukin-6 (IL-6) and C-reactive protein (CRP) concentrations (outcomes). The domains comprised metabolomic biomarkers (N = 249), proteomic biomarkers (N = 2,923), cardiometabolic conditions (N = 17), immune-mediated conditions (N = 8), infection-related traits (N = 9) and neuropsychiatric conditions (N = 7). Outcome genome-wide association studies (GWAS) included 67,428 participants for IL-6 and 204,402 for CRP. Independent genetic instruments were selected using genome-wide significance (P < 5 × 10⁻⁸), linkage disequilibrium (LD) clumping (R² < 0.001; window = 10,000 kb) and a minor allele frequency (MAF) threshold of > 0.01. Proxy single-nucleotide polymorphisms (SNPs) were used where required. Alleles were harmonised to the forward strand, and ambiguous palindromic variants were removed. For proteomic traits, only cis protein quantitative trait loci (cis-pQTLs; within ±1 Mb of the encoding gene) were used as instruments in the primary analysis. Primary MR analyses used inverse-variance weighting (IVW) for multi-variant instruments and the Wald ratio for single-variant instruments. Sensitivity analyses included the weighted median and MR-Egger methods. Steiger filtering was used to retain instruments consistent with the hypothesised exposure-to-outcome direction. In total, 1,890 exposure/outcome pairs were tested for CRP and 1,906 for IL-6. Multiple testing was addressed using Bonferroni correction (α = 0.05/N, where N is the number of exposures tested for each outcome) and false discovery rate control (FDR; q < 0.05). For CRP, 72 associations met the Bonferroni threshold and 149 met the FDR threshold; for IL-6, the corresponding numbers were 9 and 32. Bidirectional MR was performed for traits meeting the predefined significance threshold (Bonferroni for CRP; FDR for IL-6) to assess potential reverse causality. In reverse-direction analyses, no CRP associations met either threshold, while 18 IL-6 associations met the Bonferroni threshold and 20 met the FDR threshold.

### 2.1 Outcomes: Serum CRP and Serum/Plasma IL-6 Levels

The outcomes of interest were circulating IL-6 and CRP levels.

For IL-6, the GWAS from the CHARGE Inflammation Working Group was utilised. This data comprised a meta-analysis of up to 67,428 individuals of European ancestry and identified three independent genome wide significant loci associated with IL-6 levels [24]. GWAS summary statistics for CRP were obtained from the same consortium, comprising 204,402 individuals of European ancestry, and identified 58 loci associated with CRP levels [25]. While a larger CRP GWAS has been published by Said et al. [26],the complete GWAS summary statistics required for harmonisation were not publicly accessible at the time the analyses were conducted. We therefore used the widely adopted Ligthart et al. [25] GWAS, which remains commonly used in MR studies and provides a well-characterised set of genome-wide significant CRP instruments. Participant recruitment, quality control procedures, phenotype definitions and ethical approvals for each contributing GWAS are described in the original publications [24,25] and summarised in Supplementary Table 1.

### 2.2 Exposures: molecular, physiological and disease phenotypes

To ensure suitability for two sample MR and comparability across studies, exposure GWAS’s were included based on the following criteria:

1. Availability of complete summary statistics with effect estimates, standard errors, and genome wide coverage.
2. Availability of European ancestry data to minimise artifact from population stratification.
3. Presence of genome wide significant SNPs (P < 5×10⁻⁸) after clumping.
4. Relevance to possible systemic inflammation.
5. Where multiple GWASs were available for the same phenotype, the most recent and/or largest study was selected, prioritising those with the greatest sample size and statistical power.

Application of these criteria yielded 3,213 unique phenotypes. Detailed participant recruitment, eligibility criteria, quality control procedures (including assessments of between-study heterogeneity in meta-analyses where applicable), phenotype definitions, measurement procedures, and ethical approvals for each contributing GWAS are described in the original publications and summarised in Supplementary Table 2. All exposure GWAS summary statistics were obtained from publicly available sources, primarily from the original study publications where available and, where necessary, from the GWAS Catalog (https://www.ebi.ac.uk/gwas/). To ensure consistency across datasets, GWAS summary statistics which were not originally aligned to genome build 37 (GRCh37) were harmonised to GRCh37, as this was the most commonly used reference build across the included studies. Exposure and outcome summary statistics were obtained from independent published GWASs and consortia where possible, however, owing to the large number of exposures GWAS datasets included in the analysis, participant overlap between exposure and outcome studies could not be systematically assessed for every exposure-outcome pair.

### 2.3 Exposure domains

To aid interpretation across heterogeneous traits, exposures were grouped into six domains based on established literature and shared mechanistic themes:

1. Metabolomic: Meta analysis of circulating metabolites in Estonian and UK Biobank participants (European ancestry population n = 599,249; 249 exposure traits) [27].
2. Proteomic: circulating protein levels for 2,923 proteins from the Pharma Proteomics Project (UKB-PPP; n = 54,219; 2,923 exposure traits) [28].
3. Cardiometabolic: biomarkers, physiologic and disease traits (e.g. BMI, glycaemic traits, blood pressure, lipids) (17 exposure traits) [29–36].
4. Neuropsychiatric: psychiatric disorders, psychological traits, and neurobehavioral phenotypes (7 exposure traits) [37–43].
5. Immune mediated inflammatory diseases (IMIDs): autoimmune and autoinflammatory disease liabilities (8 exposure traits) [44–47].
6. Infection-related diseases: traits capturing susceptibility to, or consequences of, infectious diseases (9 exposure traits) [36,48–50].

These domains were specified before analysis and used solely as an interpretative framework; all MR analyses were performed at the individual-trait level, no data-driven clustering was applied.

All continuous exposure and outcome GWAS reported effect sizes per standard deviation (SD) increase in the trait; therefore, MR estimates reflect the effect on the outcome per SD increase in the genetically proxied exposure. For binary exposures, GWAS estimates were reported on the log-odds scale, and MR estimates should be interpreted as the effect of genetic liability to the exposure rather than the effect of the diagnosed condition itself.

### 2.4 Instrument selection for MR

For each exposure, genome wide significant SNPs (P < 5 × 10⁻⁸) were first extracted as candidate instruments. To ensure independence, variants were clumped using a pairwise linkage disequilibrium (LD) threshold of R² < 0.001 within a 10,000 kb window, where R denotes the correlation coefficient between two SNPs and R² the proportion of shared variance [51]. SNPs were clumped using the European 1000 Genomes project reference panel matched to the corresponding genome build [52].

If an exposure SNP was absent from the outcome GWAS, proxy variants were identified in the outcome dataset using an LD threshold of R² > 0.8; from these, the proxy with the highest R² that was also present in the exposure dataset was selected.

Following instrument selection, SNPs were harmonised with outcome datasets by aligning alleles to the forward DNA strand; ambiguous palindromic variants with unresolved strand orientation were excluded. The final number of SNPs contributing to each exposure outcome instrument set is reported in Supplementary Tables 3 and 4.

### 2.5 MR instruments for plasma proteomic exposures (cis-pQTLs)

For plasma proteomics, instruments were categorised as *cis*- or *trans*- pQTLs (protein quantitative trait loci) [53]. SNPs located within ±1 megabase (Mb) of the gene encoding the respective protein were classified as *cis*-pQTLs, while those outside this window (or on different chromosomes) were classified as *trans*-pQTLs. SNPs in the *cis* region of the gene are more likely to influence protein abundance through regulation of the encoding gene and are therefore generally considered less susceptible to horizontal pleiotropy than *trans*-pQTLs [54,55]. Consequently, primary analysis was restricted to *cis*-pQTLs instruments, while analyses including *trans-*pQTLs were performed as sensitivity analysis. Gene coordinates were obtained from the Olink/UKB-PPP proteomics metadata, and SNP genomic positions were matched using the pQTL summary statistics [28].

### 2.6 Mendelian randomisation (MR) methods

Analyses were conducted using a two-sample Mendelian randomisation (MR) framework and were structured into primary and sensitivity analysis components. Prior to MR estimation, variant directionality was assessed using Steiger filtering, which compares the variance explained by each genetic variant in the exposure versus the outcome to confirm the assumed causal direction [56]. For proteomic traits, only *cis*-acting instruments were included in the primary analyses, as described above.

Causal effects were estimated using standard MR approaches, with the choice of estimator determined by the number of available genetic instruments for each exposure outcome pair. When a single genetic instrument was available, causal effects were estimated using the Wald ratio. When two instruments were available, effects were estimated using the inverse variance weighted (IVW) method. When three or more instruments were available, IVW was used as the primary estimator, with weighted median [57] and MR-Egger [19] performed as supplementary sensitivity analyses. This strategy was implemented as MR-Egger regression and its associated pleiotropy diagnostics are not well defined for fewer than three instruments [19], and weighted median estimates provide limited additional robustness when very few instruments are available [32]. The weighted median estimator provides consistent causal estimates under the assumption that at least 50% of the total instrument weight derives from valid instruments, while MR-Egger regression allows for directional horizontal pleiotropy under the Instrument Strength Independent of Direct Effect (InSIDE) assumption, whereby the strength of the genetic instrument-exposure association is independent of its direct pleiotropic effect on the outcome [19,57]. Strength of the instrumental variables was assessed using F statistics (Supplementary Table 3 and 3). As is standard in the MR literature, F statistics greater than 10 were considered indicative of sufficiently strong instruments and reduced risk of weak instrument bias [58].

Sensitivity analyses were conducted to assess the robustness of findings and the validity of MR assumptions. Steiger filtering was applied prior to MR estimation to remove variants more strongly associated with the outcome than the exposure. Instrument strength was evaluated using F-statistics to assess susceptibility to weak instrument bias. Directional horizontal pleiotropy was evaluated using the MR-Egger intercept test, and heterogeneity among SNP-specific causal estimates was assessed using Cochran’s Q statistic.

Multiple testing correction was applied separately for each outcome and MR estimator using standard Bonferroni and false discovery rate (FDR) procedures.

### 2.7 Bidirectional Mendelian randomisation analysis

To evaluate whether potential causal effects identified in the primary analyses (trait to IL-6/CRP) might also reflect causal effects in the reverse direction (IL-6/CRP to trait), we performed bidirectional MR analyses using IL-6 and CRP as exposures. As outcomes we focused on traits that had evidence of possible causal effects on IL-6/CRP in the primary analyses after multiple testing correction (FDR for IL-6; Bonferroni for CRP). For IL-6, the Bonferroni-corrected threshold yielded very few eligible traits, limiting the ability to meaningfully assess potential bidirectional effects. Therefore, we used the more permissive FDR threshold to avoid overlooking plausible associations while still controlling for false discovery.

Bidirectional IL-6 and CRP instruments were restricted to respective *cis*-pQTLs for primary analysis, using the same parameters as applied to the proteomic traits in the primary analysis (SNPs within ±1 Mb of gene). Due to the lack of *cis* variants for circulating IL-6 within the *IL-C* gene, variants within the IL-6 receptor gene (*ILCR*) were used as proxies for IL-6 signalling in line with existing literature [59–61]. All other parameters for instrumentation were same as forward analysis.

### 2.8 Multivariable Mendelian randomisation (MVMR) and Mediation analysis

We examined the effects of IL-6 and BMI on CRP levels using MVMR and MR mediation [62,63]. Using standard techniques for MVMR and MR mediation [63], IVs for BMI and IL-6 were combined and harmonised with CRP summary statistics using the same procedures defined above for univariable MR analyses. Independent variants associated with either exposure were retained, and effects were estimated using IVW method in MVMR. This approach estimates the direct effect of each exposure (IL-6 and BMI) on the outcome (CRP) while accounting for the other exposure in the model.

To assess mediation (whether BMI influences CRP via IL-6), we applied a two-step MR framework. Causal effects from were estimated using the above procedures. The causal effect of BMI on IL-6 was estimated. Second, the effect of IL-6 on CRP was estimated. The indirect effect of BMI on CRP through IL-6 was calculated as the product of these two estimates (β BMI →IL-6 × β IL-6 →CRP). The total effect of BMI on CRP was obtained from univariable MR, and the direct effect was derived by subtracting the indirect effect from the total effect. The proportion mediated was calculated as the ratio of the indirect effect to the total effect. Standard errors for the indirect effect were estimated using the delta method.

### 2.9 Evidence appraisal across MR methods

In addition to sensitivity analyses described above, an evidence appraisal framework was applied to assess the robustness of findings across complementary MR approaches. MR findings were considered *robust* if all of the following criteria were met. In the manuscript such findings are mentioned as “Robust across MR Methods”:

1. The IVW estimate and at least one sensitivity method (weighted median or MR-Egger) were directionally concordant.
2. The supporting sensitivity estimate survived false discovery rate correction (FDR < 5%).
3. For MR-Egger support, the intercept test indicated no evidence of directional horizontal pleiotropy (P > 0.05).

Robustness could not be evaluated in exposure outcome pairs instrumented by fewer than three variants as these are not suitable for multiple MR methods or MR-Egger intercept tests.

### 2.10 Availability of data and code

All GWAS summary statistics used in this study were publicly available from the cited sources; links to which can be found in Supplementary Tables 1 and 2. Code and scripts used for data processing and analysis are available at DeterminingSystemicInflammation (GitHub).

All analyses were performed in Rstudio (version 4.6.0). The TwoSampleMR package (Version 0.7.5) [64] was used for Mendelian randomisation analyses. Linkage disequilibrium clumping and proxy variant identification were conducted using the ieugwasr package (version 1.1.0) [65] and the European 1000 Genomes Project reference panel. Additional data processing and statistical analyses were performed using standard R packages as detailed in Supplementary Table 6.

This study was reported in accordance with the Strengthening the Reporting of Observational Studies in Epidemiology using Mendelian Randomisation (STROBE-MR) guidelines [66]. The completed STROBE-MR checklist is available as part of the extended data deposited on Figshare [22].

## 3. Results

### 3.1 Summary

After Bonferroni correction, we identified 72 traits with potential causal effects on CRP levels (Table 1, Figure 2) and 9 traits with potential causal effects on IL-6 levels (Table 2, Figure 3). After FDR correction potential causal effects were observed for 149 traits on CRP and 32 traits on IL-6 (Supplementary Table 3). Sensitivity analyses using the weighted median and MR-Egger methods, to assess the influence of directional pleiotropy and potential invalid instruments, are presented in Supplementary Tables 3. Analysis including cis-pQTL instruments are presented in Supplementary Table 3.

**Figure 2.**
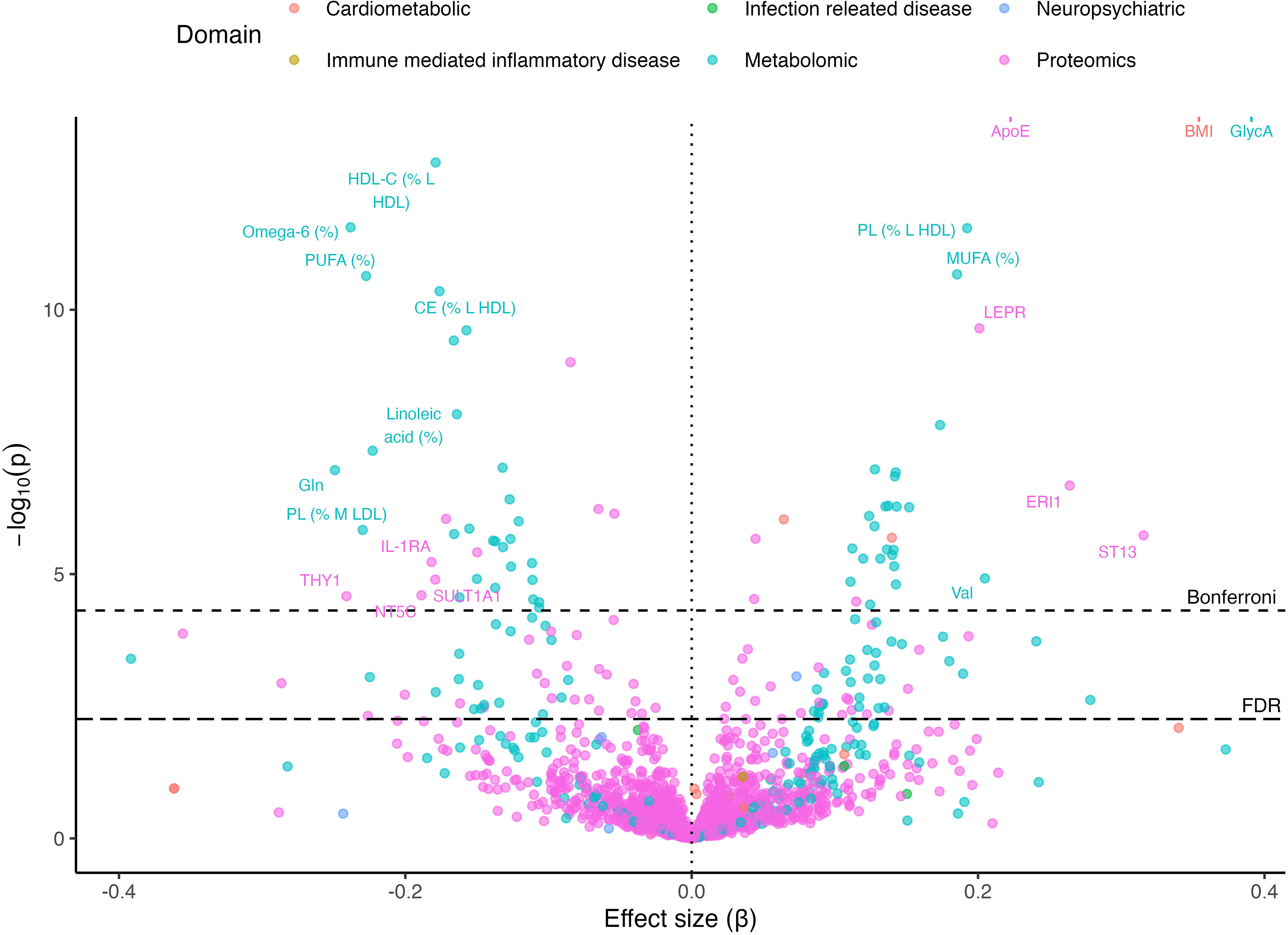
Volcano plot of MR effect estimates for molecular, physiologic and disease traits on circulating CRP. The x-axis shows the causal effect estimate (β), expressed as the change in CRP (in SD units) per SD increase in the genetically proxied exposure. The y-axis shows the -log₁₀(P) value. Analyses were restricted to Steiger-consistent results, primary MR methods only (inverse weighted or Wald ratio), and cis-only instruments for proteomic exposures. The vertical dotted line denotes the null (β = 0). The horizontal dashed lines indicate the Bonferroni (4.88 × 10⁻⁵) (small dashes) and FDR (5.52 × 10⁻³) (large dashes) thresholds. Of the 72 Bonferroni-significant causal effects reported in Table 1, only the 20 exposures with the largest absolute effect sizes are labelled to improve readability. For visualisation purposes, the plotted -log₁₀(P) values for body mass index (BMI), obesity, and Glycoprotein acetyls (GlycA) were capped at 13, although their true P values were 5. 55 x 10-70, 3.59 x 10-36 and 6.99 x 10-19, respectively. Colours indicate the domain of each exposure.

**Figure 3.**
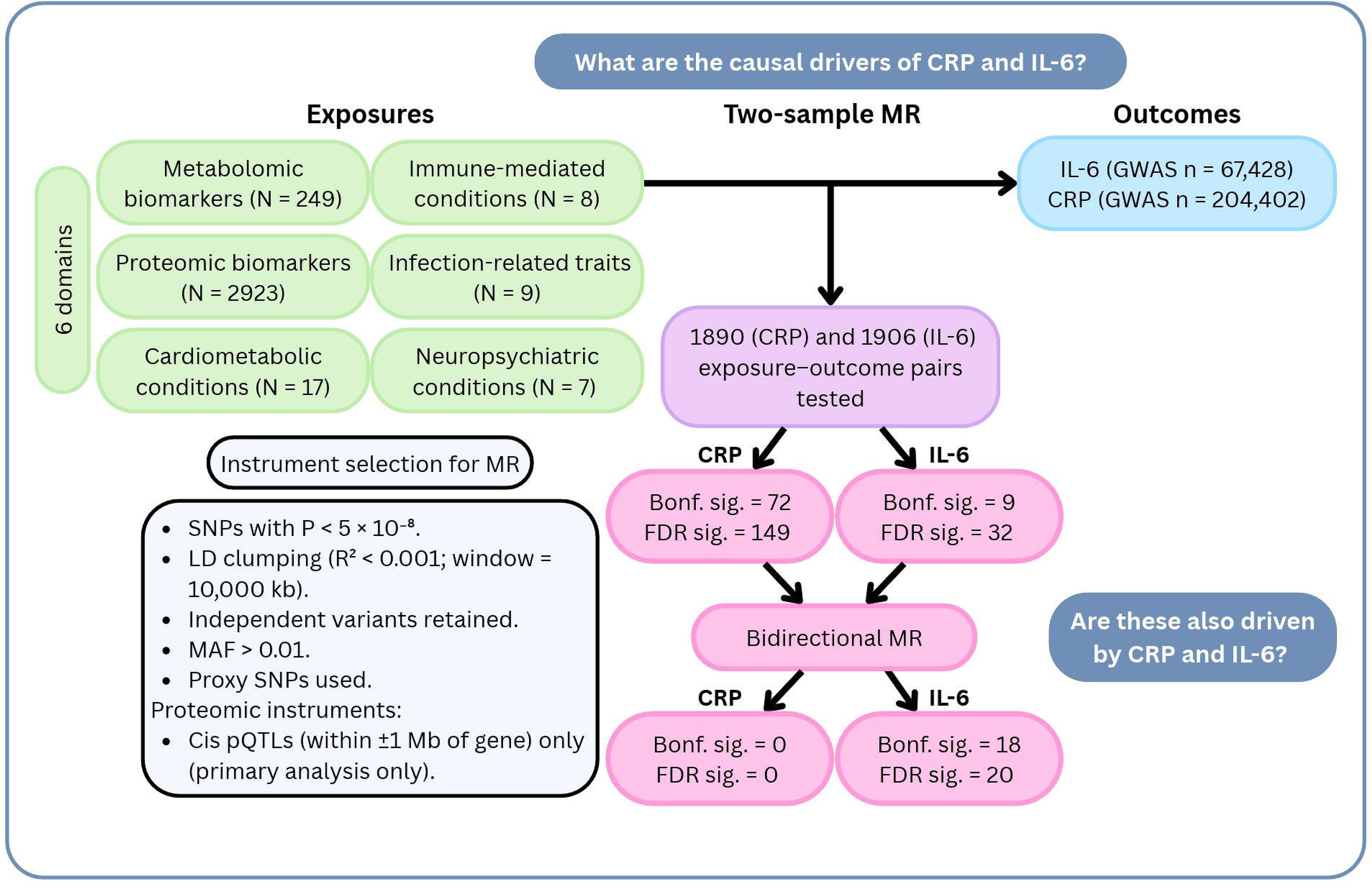
Volcano plot of MR effect estimates for molecular, physiologic and disease traits on circulating IL-6. The x-axis shows the causal effect estimate (β), expressed as the change in IL-6 (in SD units) per SD increase in the genetically proxied exposure. The y-axis shows the -log₁₀(P) value. Analyses were restricted to Steiger-consistent results, primary MR methods only (inverse weighted or Wald ratio), and cis-only instruments for proteomic exposures. The vertical dotted line denotes the null (β = 0). The horizontal dashed lines indicate the Bonferroni (4.77 × 10⁻⁵) (small dashes) and FDR (1.52 × 10⁻³) (large dashes) thresholds. Exposures labelled in the plot met the Bonferroni-corrected significance threshold. For visualisation purposes, the plotted -log₁₀(P) values for obesity and body mass index (BMI) were capped at 6, although their true P values were 4.94× 10⁻14, 3.58× 10⁻22, respectively.

**Table 1.**
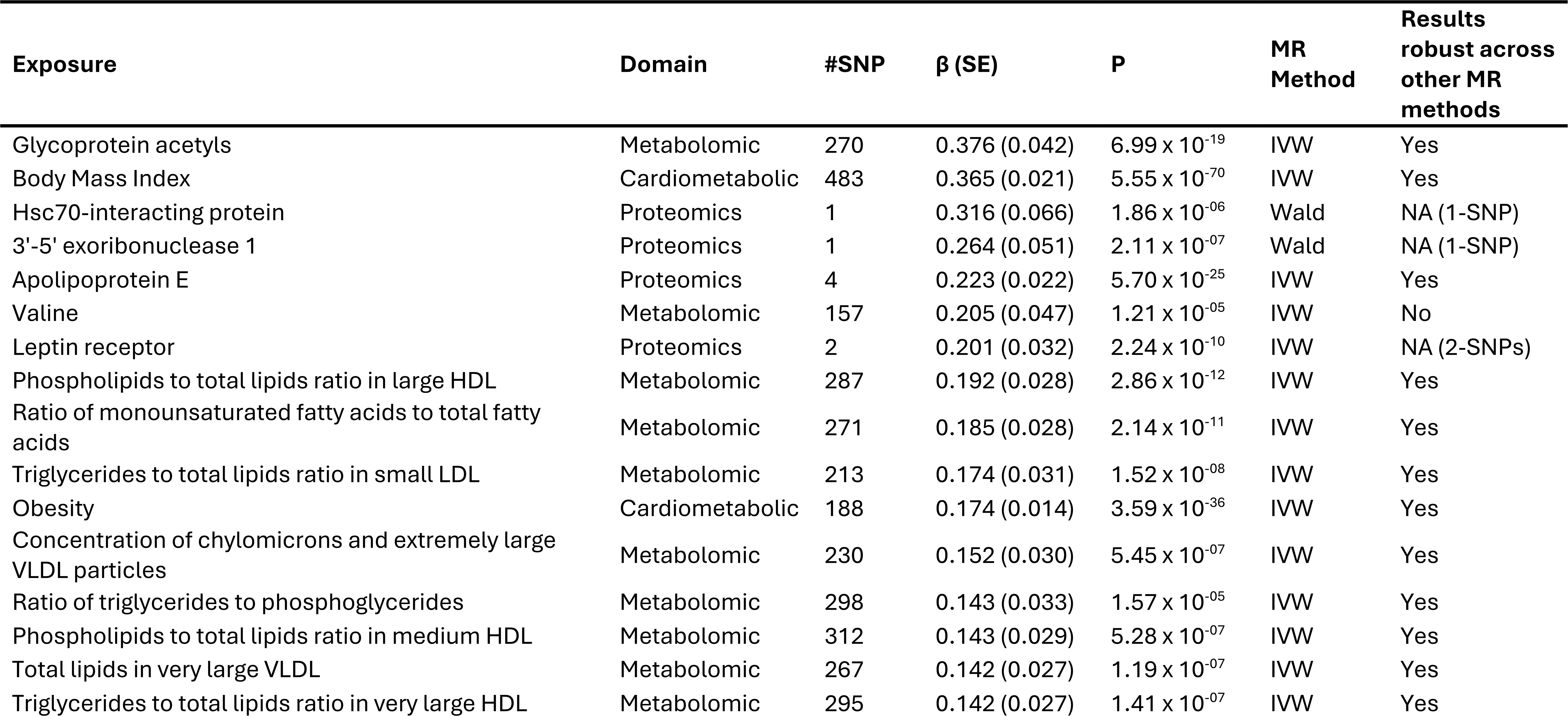
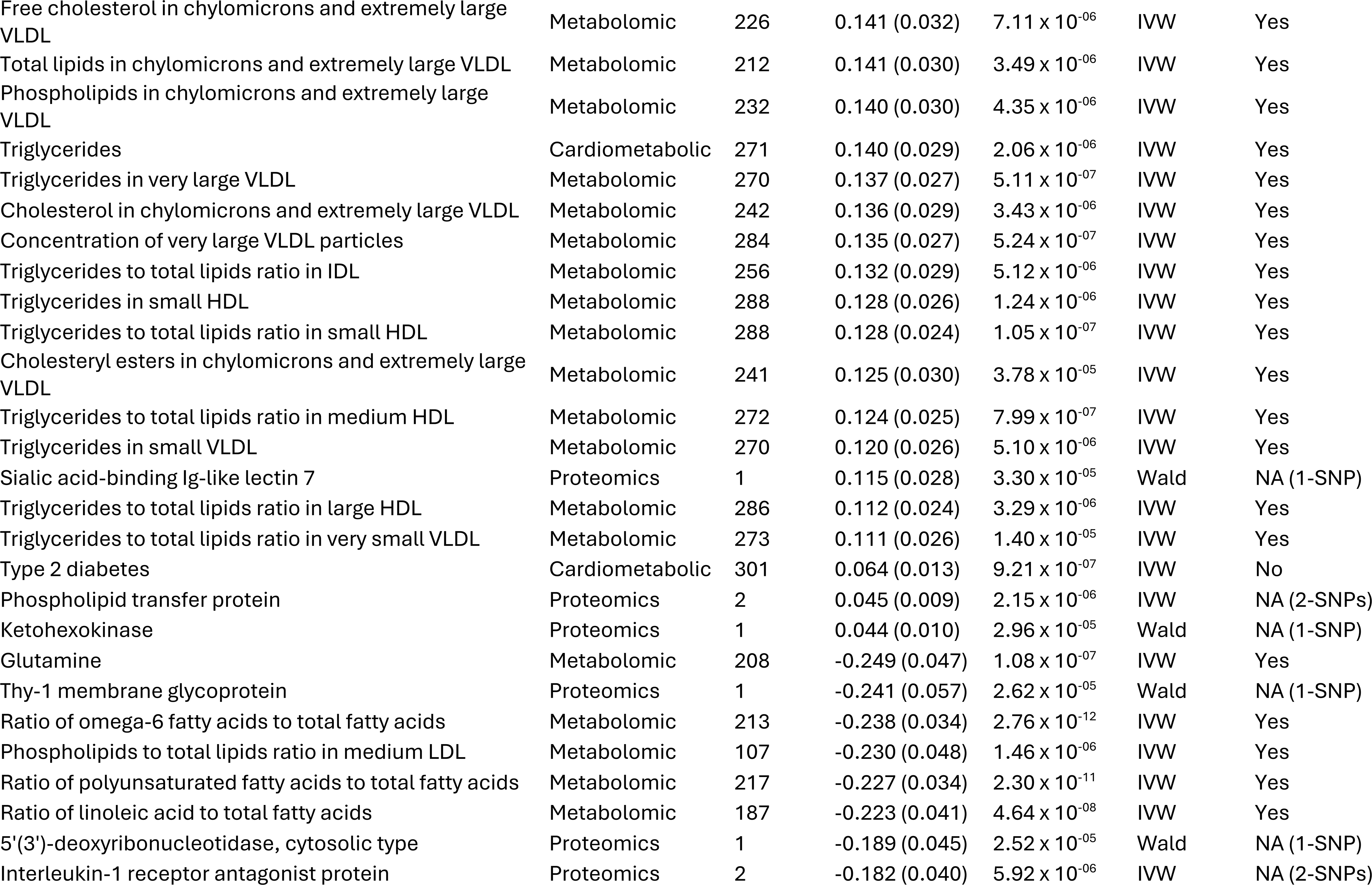
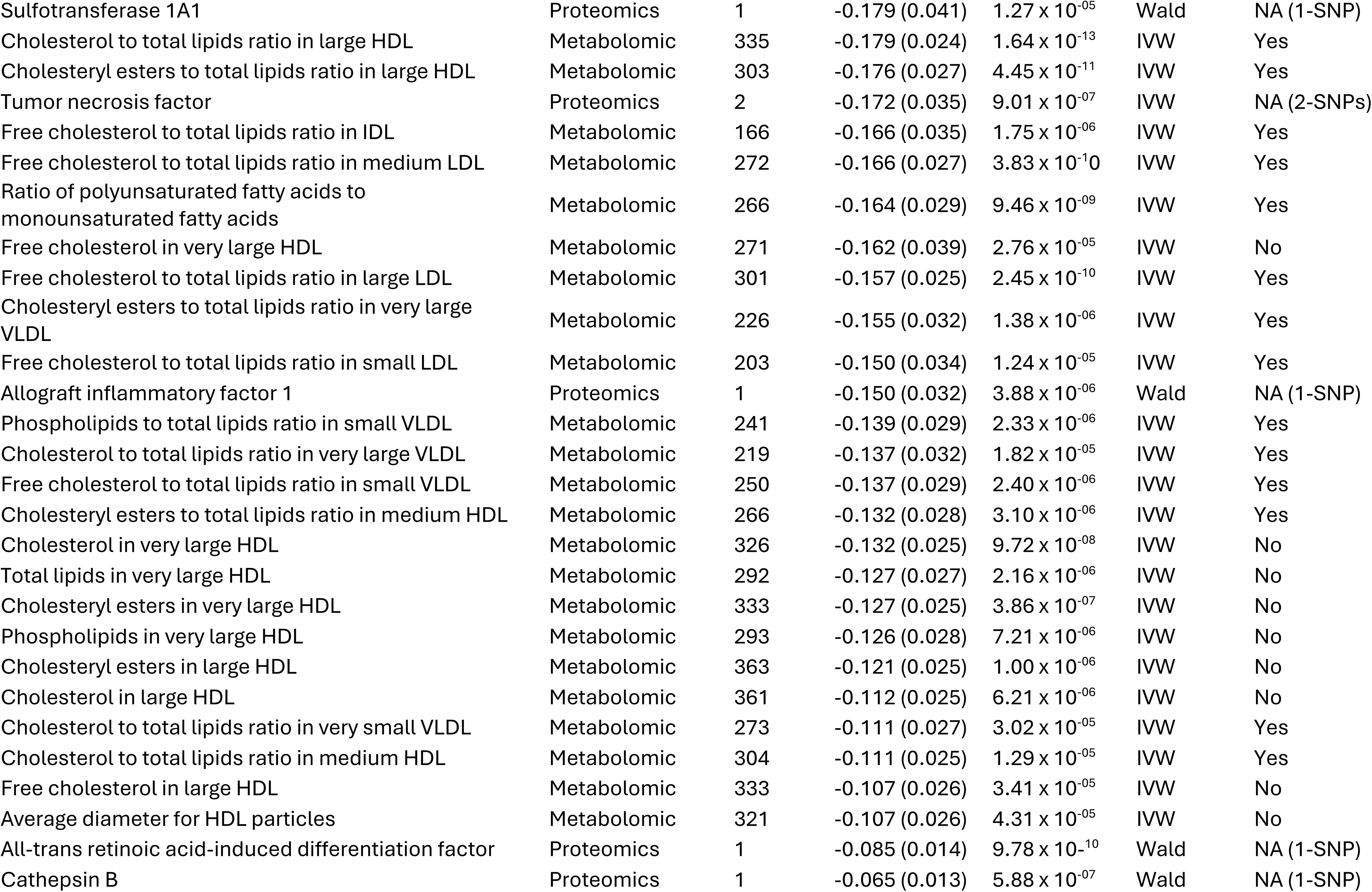
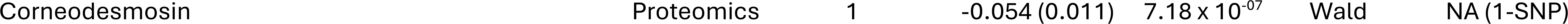
Mendelian randomisation results showing traits with evidence of potential causal effects on circulating CRP levels after Bonferroni correction. Estimates represent the effect of each genetically proxied exposure on CRP, expressed as the change in CRP (in SD units) per SD increase in the exposure (for continuous traits) or per unit increase in genetic liability (for binary traits). Results are from the primary MR methods (inverse variance weighted or Wald ratio). Results robust across other MR methods was assessed only for analyses with ≥3 instruments, requiring directionally concordant IVW, weighted median, and MR-Egger estimates robust to FDR correction and a non-significant MR-Egger intercept test; for analyses with fewer instruments, this assessment was not applicable. Bonferroni and FDR thresholds were 4.88 × 10⁻⁵ and 5.52 × 10⁻³, respectively. Results robust across other MR methods is represented as NA when the analysis does not have enough SNPs for all supplementary methods.

### 3.2 Causal drivers of circulating CRP

Of the 72 traits showing evidence of potential causal effects on circulating CRP levels after Bonferroni correction (Table 1), the majority were metabolomic traits (n = 52; 72%), along with several proteomic traits (n = 16; 22%) and a few cardiometabolic traits (n = 4; 6%). Among metabolomic traits, potential causal drivers of CRP were dominated by lipid and fatty acid-related measures. Collectively, these findings indicate that genetically proxied increases in amino acids such as valine, triglyceride rich lipoproteins, adiposity, and selected circulating proteins increase CRP levels, whereas higher genetically predicted levels of polyunsaturated fatty acid-related traits, glutamine, and multiple HDL-associated lipid measures decrease CRP.

Higher genetically predicted levels of Glycoprotein acetyls (GlycA; β = 0.376, SE = 0.042, P = 6.99 × 10⁻¹⁹), the ratio of monounsaturated fatty acids to total fatty acids (β = 0.185, SE = 0.028, P = 2.14 × 10⁻^11^), and multiple triglyceride rich lipoprotein measures, including triglycerides in small and very large VLDL particles and triglyceride to-total lipid ratios across LDL and HDL subclasses, increase CRP levels. These findings were robust across methods. In contrast, higher genetically predicted levels of HDL-related cholesterol measures and polyunsaturated fatty acid-related traits decrease CRP levels, including the cholesterol-to-total lipids ratio in large HDL (β = -0.179, SE = 0.024, P = 1.64 × 10⁻¹³), the ratio of polyunsaturated fatty acids to total fatty acids (β = -0.227, SE = 0.034, P = 2.30 × 10⁻¹¹), and related PUFA and HDL subclass measures. Higher genetically predicted glutamine levels also decrease CRP (β = -0.249, SE = 0.047, P = 1.08 × 10⁻⁷). Effect sizes were moderate and displayed coherent scaling across related metabolic traits, consistent with a structured metabolic perturbation.

Of the cardiometabolic traits, higher genetically predicted body mass index (β = 0.365, SE = 0.021, P = 5.55 × 10⁻⁷⁰), obesity (β = 0.174, SE = 0.014, P = 3.59 × 10⁻³⁶) and Triglycerides (β = 0.140, SE = 0.029, P = 2.06 × 10⁻⁶) increased CRP. These effects were robust across methods. Type 2 diabetes liability (β = 0.064, SE = 0.013, P = 9.21 × 10⁻⁷) also increased CRP levels, however this effect was not robust due lack of evidence across methods.

Higher genetically predicted levels of Apolipoprotein E (β = 0.223, SE = 0.022, P = 5.70 × 10⁻²⁵) increased CRP; these effects were robust across methods. Several additional proteins showed potential causal effects on CRP (decreasing), including Thy-1 membrane glycoprotein (β = -0.241, SE = 0.057, P = 2.62 × 10⁻⁵), tumour necrosis factor (β = -0.172, SE = 0.035, P = 9.01 × 10⁻⁷), and interleukin-1 receptor antagonist protein (β = -0.182, SE = 0.040, P = 5.92 × 10⁻⁶). However, these exposures were instrumented by one or two genetic variants only. While these variants may represent strong instruments, sensitivity analyses using alternative MR estimators and formal tests for horizontal pleiotropy could not be performed due to limited number of genetic variants available, and these results should be interpreted with caution.

### 3.3 Causal drivers of circulating IL-6

Primarily cardiometabolic and lipid-related traits showed evidence of causal effects on circulating IL-6 levels after Bonferroni correction (Table 2). Collectively, the findings suggest that genetically proxied adiposity increase circulating IL-6, while higher HDL-related lipid measures and IL-1 receptor antagonist levels decrease IL-6, although evidence for robustness across MR methods was limited.

**Table 2.**
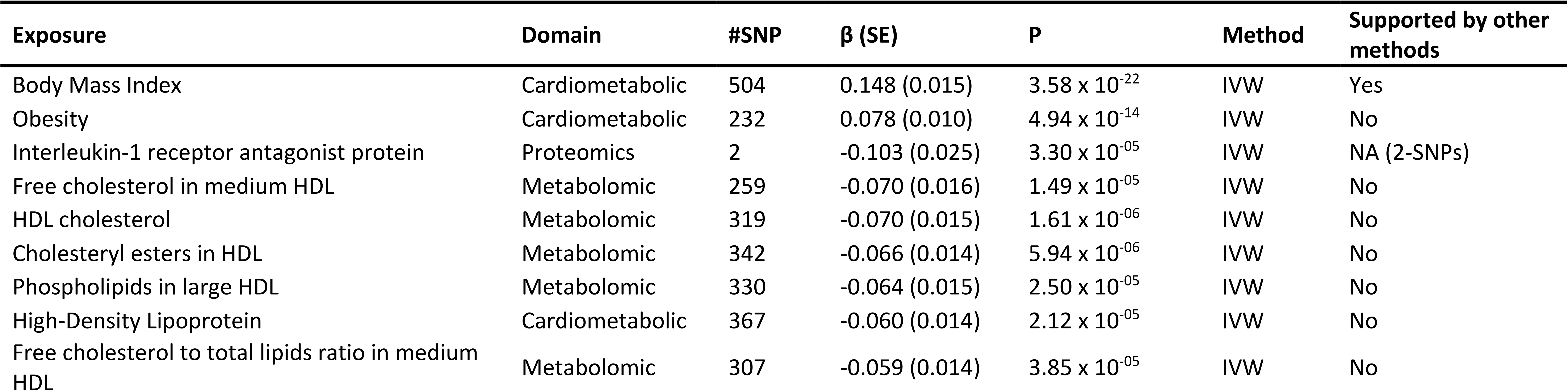
Mendelian randomisation results showing traits with evidence of potential causal effects on circulating IL-6 levels after Bonferroni correction. Estimates represent the effect of each genetically proxied exposure on IL6, expressed as the change in IL-6 (in SD units) per SD increase in the exposure (for continuous traits) or per unit increase in genetic liability (for binary traits). Results are from the primary MR methods (inverse variance weighted or Wald ratio). Results robust across other MR methods was assessed only for analyses with ≥3 instruments, requiring directionally concordant IVW, weighted median, and MR-Egger estimates robust to FDR correction and a non-significant MR-Egger intercept test; for analyses with fewer instruments, this assessment was not applicable. Bonferroni and FDR thresholds were 4.77 × 10⁻⁵ and 1.52 × 10⁻³, respectively. Results robust across other MR methods is represented as NA when the analysis does not have enough SNPs for all supplementary methods.

Body mass index (BMI; β = 0.148, SE = 0.015, P = 3.58 × 10⁻²²) and obesity liability (β = 0.078, SE = 0.010, P = 4.94 × 10⁻¹⁴), both increase circulating IL-6 levels, consistent with their effects on CRP. Among metabolomic traits, several high-density lipoprotein (HDL) related lipid measures decrease IL-6 (free cholesterol in medium HDL; (β = -0.070, SE = 0.016, P = 1.49 × 10⁻⁵), total HDL cholesterol; (β = -0.070, SE = 0.015, P = 1.61 × 10⁻⁶), cholesteryl esters in HDL (β = -0.066, SE = 0.014, P = 5.94 × 10⁻⁶). Consistent IL-6 lowering effects were also observed for the free cholesterol-to-total lipids ratio in medium HDL (β = -0.059, SE = 0.014, P = 3.85 × 10⁻⁵). Related cardiometabolic measures of high-density lipoprotein showed a similar direction of effect (β = -0.060, SE = 0.014, P = 2.12 × 10⁻⁵). Among proteomic traits, higher genetically predicted levels of interleukin-1 receptor antagonist decrease circulating IL-6 (β = -0.103, SE = 0.025, P = 3.30 × 10⁻⁵).

Effect estimates for IL-6 were generally modest in magnitude. Adiposity-related traits showed moderate IL-6 increasing effects (e.g., BMI β=0.148; obesity β=0.078), while HDL-related lipid measures showed consistent IL-6 lowering effects with similar effect sizes, suggesting quantitative coherence across related phenotypes rather than isolated outliers. Across IL-6 analyses, only BMI, of the Bonferroni-significant findings, met the criteria for support across multiple MR methods, reflecting either limited instrument numbers or a lack of consistency across sensitivity analyses.

In the forward analyses of both CRP and IL-6, BMP and activin membrane-bound inhibitor homolog was instrumented by a single cis-acting SNP with weak instrument strength (F = 2.83). The analysis was retained but interpreted with caution because of the potential for weak instrument bias.

### 3.4 Results from Bi-directional MR analyses

#### 3.4.1 Causal effects of CRP on metabolic traits

To explore reverse causation, bidirectional MR analyses were performed using circulating CRP as the exposure. When restricting CRP instruments to *cis*-acting variants (n = 2), no significant findings were observed. Using both *cis*- and *trans*-instruments, CRP demonstrated potential causal effects on several metabolomic traits after multiple testing correction (Supplementary Table 4). Collectively, these findings may suggest that elevated circulating CRP could have downstream effects on amino acid metabolism and lipoprotein composition, particularly HDL- and VLDL-related lipid fractions. However, these findings should be interpreted cautiously. While analyses using all available or *trans*-only instruments were supported by strong instrument strength (mean F-statistics >120), the *cis*-only analyses were based on only two variants, likely limiting statistical power and precision. Consequently, the absence of significant associations in *cis*-only analyses may reflect limited power rather than definitive evidence against a causal effect, however, it may also indicate that the observed bidirectional associations are influenced predominantly by *trans*-acting variants. Using all available instruments, higher genetically predicted CRP levels lower circulating glutamine levels (β = −0.145, SE = 0.042, P = 6.61 × 10⁻⁴), consistent with the opposite effect observed in the forward direction. CRP was also found to increase levels of glycoprotein acetyls (GlycA; β = 0.203, SE = 0.055, P = 2.35 × 10⁻⁴), suggesting a reinforcing relationship between CRP and this composite marker of systemic inflammation. In addition, higher CRP levels also increase phospholipid-to-total lipid ratios in large HDL particles (β = 0.202, SE = 0.044, P = 4.58 × 10⁻⁶).

Several inverse effects were also observed between CRP and lipid traits. Higher genetically predicted CRP levels lower phospholipid-to-total lipid ratios in small VLDL particles (β = −0.244, SE = 0.072, P = 6.73 × 10⁻⁴) and lower free cholesterol levels in very large HDL particles (β = −0.196, SE = 0.056, P = 4.43 × 10⁻⁴).

#### 3.4.2 Causal effects of IL-6 on metabolic traits

In the reverse direction, *cis*-pQTL for IL-6 showed evidence of downstream effects on lipid and lipoprotein metabolism. Higher IL-6 was associated with higher concentrations of HDL particles (β = 0.131, SE = 0.018, P = 6.97 × 10⁻^13^) and multiple HDL-related lipid fractions, including (not limited to) HDL cholesterol (β = 0.069, SE = 0.017, P = 4.11 × 10⁻^05^), cholesteryl esters (β = 0.070, SE = 0.017, P = 3.83 × 10⁻^05^) and free cholesterol (β = 0.065, SE = 0.017, P = 1.18 × 10⁻⁴).

While these findings appear directionally inconsistent with forward MR results, where higher HDL-related measures were associated with lower IL-6 levels, this may reflect the distinct biological mechanisms captured in each direction. The IL-6 instrument (rs4537545) used for this analysis is located within the *IL-CR* locus, and it is in strong LD with *IL-CR* variant rs2228145 which is known to impair IL-6 signalling [67]. Therefore, the findings observed here are consistent with impaired *IL-CR* signalling being associated with higher HDL levels. As such, the bidirectional MR results may reflect downstream effects of altered IL-6 signalling on lipid metabolism, rather than a direct effect of IL-6 concentration *per se*. Furthermore, this analysis relied on a single *cis*-acting instrument. Although the instrument was strong (F = 397.7), analyses based on a single variant limit the ability to evaluate robustness across complementary MR methods.

### 3.5 Effects of IL-6 and BMI on CRP

Using the above *cis*-pQTL for IL-6, we examined the effect of IL-6 on CRP. This analysis was based one SNP in *IL-CR* (rs4537545), which showed a large negative effect on CRP (β = −0.93, SE = 0.04, P = 5.51 × 10⁻¹²³). As described above, rs4537545 is in strong LD (r² = 0.95) [25] with the *IL-CR* variant rs2228145 which is known to lower CRP levels by impairing IL-6 receptor expression and signalling [67]. Thus, our results indicate that *IL-CR* variant known to impair IL-6 signalling lowers CRP levels.

Given the strong effects of IL-6 and BMI on CRP, and it is well known that adipose cells produce IL-6 [68], we conducted MR mediation and MVMR analyses to examine whether higher BMI increases CRP levels by influencing IL-6 levels (i.e., BMI → IL-6 → CRP). Two-step MR mediation analysis (Supplementary Table 5b) showed a modest inverse indirect effect of BMI on CRP through IL-6 (β indirect = −0.14, SE = 0.01, P = 5. 8 x 10 ^-19^), alongside a larger positive direct effect of BMI on CRP (β direct = 0. 50, SE = 0.026, P = 8.47 x 10 ^-82^). These findings suggest that although BMI influences both IL-6 and CRP, the positive effect of BMI on CRP is unlikely to be substantially mediated through IL-6.

MVMR analyses were undertaken (Supplementary Table 5c). However, there was a marked imbalance in genetic instruments available for MVMR between BMI (462 variants) and IL-6 (only one variant; rs4537545). Additionally, conditional F statistics were calculated for the MVMR analyses to assess instrument strength after accounting for the correlation between exposures (Supplementary Table 5d). While BMI retained adequate conditional instrument strength (F = 14.75), IL-6 showed very weak conditional instrument strength (F = 1.97). Consequently, the IL-6 estimate is susceptible to weak instrument bias (under-estimating the IL-6 effect) and the subsequent results difficult to interpret.

## 4. Discussion

### 4.1 Summary

In this study we have systematically examined potential causal drivers of systemic inflammation by testing potential causal effects of >3000 molecular, physiologic and disease traits on circulating CRP and IL-6 levels. Our work identified multiple causal determinants of circulating IL-6 and CRP, with the strongest evidence observed within metabolomic, metabolic and proteomic traits. Effect directions were largely consistent across sensitivity analyses, supporting the robustness of the primary findings.

### 4.2 CRP vs IL6: differences and similarities

Although IL-6 and CRP are closely linked components of the inflammatory cascade and are often interpreted together, our findings highlight important differences in their causal relationships with metabolic traits. Adiposity-related traits emerged as shared upstream determinants of both biomarkers. However, effect sizes were substantially larger and more robust for CRP than for IL-6. In particular, BMI, alongside obesity and T2D, showed the largest effects on CRP consistent with extensive observational and genetic evidence positioning CRP as a downstream integrator of systemic metabolic and inflammatory disturbances [69].

In contrast to CRP, IL-6 levels are influenced by fewer traits, and the magnitude of these effects are more modest. Notably, multiple biologically related HDL traits decrease IL-6 levels, suggesting quantitative coherence rather than isolated or spurious associations. This pattern is consistent with the more complex regulation of IL-6 and signalling pathways [70]. However, BMI was the only trait to be robust across MR methods, despite most traits having enough genetic instruments for assessment.

Nevertheless, in addition to biological differences, the results may also be driven by be methodological differences in the underlying GWAS datasets. The CRP GWAS was substantially larger than the IL-6 GWAS and yielded a greater number of associated loci, resulting in stronger genetic instruments and increased statistical power. In addition, differences in trait heritability, measurement characteristics and instrument availability may have contributed to the greater number of significant findings observed for CRP. Consequently, some of the apparent asymmetry between IL-6 and CRP may reflect such differences rather than true biological differences alone.

### 4.3 Adiposity as a shared upstream driver of inflammation

Given the well-established role of IL-6 signalling in stimulating hepatic CRP production [71], we explored whether if effect of adiposity on CRP is mediated through IL-6. Consistent with previous studies [24], BMI showed strong positive effects on both circulating IL-6 and CRP. However, there was little evidence to support IL-6 as a substantial mediator of the effect of BMI on CRP. Two-step MR mediation analyses provided little evidence for an indirect effect of BMI on CRP via IL-6, and in MVMR, the interpretation of the IL-6 estimate is limited by weak conditional instrument strength, likely driven by the single IL-6 instrument.

The available genetic instruments for circulating IL-6 are limited (n = 1) and capture complex biological mechanisms. The variant used, rs4537545, is located in the IL6R locus [25,67] and is in strong LD with the functional *IL-CR* variant rs2228145 (Asp358Ala) which is associated a 2-fold increase in soluble IL-6R (sIL-6R) serum levels resulting in reduced IL-6-induced CRP [72]. As such, this instrument may proxy altered IL-6 signalling rather than IL-6 activity itself, complicating causal interpretation. Due to the singular IL-6 instrument, sensitivity analyses such as MR-Egger regression, weighted median estimation, and heterogeneity testing could not be performed. Therefore, IL-6-exposure analyses rely heavily on the validity of this biologically characterised instrument and should be interpreted with appropriate caution.

Taken together, these findings suggest that although adiposity influences both IL-6 and CRP levels, the effect of BMI on CRP may operate largely through pathways independent of circulating IL-6, or through inflammatory mechanisms not adequately captured by current IL-6 genetic instruments.

Recent methodological work has highlighted that genetic liability to CRP may partially capture adiposity-related pathways [73]. Consequently, the use of CRP instruments, particularly *trans*-acting variants, may bias results due to their correlation with BMI [73]. We observed associations suggesting that higher circulating CRP may influence amino acid metabolism and lipoprotein composition; however, these effects were driven primarily by *trans*-acting variants. Therefore, these results should be interpreted with caution, as the observed signal may reflect pleiotropic effects of adiposity-related pathways rather than direct causal effects of CRP.

### 4.4 Inflammation and lipid metabolism

Lipid metabolism emerged as a key point of convergence for both biomarkers. HDL, traditionally considered cardioprotective [74], represents a heterogeneous group of particles whose composition and function may be differentially influenced by inflammatory signalling [74]. Earlier observational work largely focussed on total HDL cholesterol as a protective marker, suggesting an inverse relationship between HDL levels and systemic inflammation [75]. However, more recent studies have highlighted HDL heterogeneity and functional alterations, demonstrating that inflammation can remodel HDL particles [76] and push it toward a more ‘pro inflammatory’ phenotype during systemic inflammation. In our primary analyses, higher HDL-related measures were associated with lower circulating IL-6. In contrast, triglyceride rich and VLDL-related lipid measures were associated with increased CRP, whereas HDL cholesterol and polyunsaturated fatty acid profiles were associated with lower CRP. These findings align with experimental and clinical evidence linking inflammation to altered lipoprotein metabolism and HDL remodelling [77]. Moreover, reverse MR analyses demonstrated strong and consistent effects of IL-6 on multiple HDL-related traits, supporting a model in which IL-6 exerts clearer downstream metabolic consequences than can be detected for its upstream determinants.

Proteomic exposures further highlighted plausible inflammatory regulators, including IL-1 receptor antagonist (IL-6 and CRP), apolipoprotein E (CRP only), and the leptin receptor (CRP only). While these findings are biologically consistent with established inflammatory and metabolic pathways [67], many were based on limited number of genetic instruments and should therefore be interpreted cautiously given reduced estimator stability and limited capacity to assess horizontal pleiotropy [19].

Taken together, these results support a model in which adiposity acts as a shared upstream driver of low-grade systemic inflammation, captured more strongly by CRP, while the effects on IL-6 are more constrained and perhaps context-dependent within the inflammatory–metabolic network. CRP appears to function primarily as a broad readout of cumulative inflammatory burden, whereas IL-6 results demonstrate a narrower set of causal drivers alongside downstream effects largely related to lipid metabolism. This distinction underscores the importance of considering IL-6 and CRP as complementary, but not interchangeable, biomarkers of inflammation.

IL-6 signalling is biologically complex, operating through multiple pathways including classic and trans signalling, which can have distinct – and in some contexts opposing – effects [70]. Classic signalling is often homeostatic and anti-inflammatory, whereas trans signalling is typically pro-inflammatory [70]. Circulating IL-6 concentrations, as captured in GWAS, represent a composite biomarker that does not distinguish between these signalling modes or their tissue specific contexts.

As a result, genetically predicted differences in upstream traits may influence specific components of IL-6 biology without substantially altering total circulating IL-6 levels. This could attenuate detectable causal estimates when IL-6 is analysed as a single composite outcome, potentially contributing to the modest effect sizes and limited support observed. Additionally, the limited number of genetic instruments available for IL-6 may have reduced statistical power to detect causal effects. In contrast, CRP represents a downstream acute phase response integrating IL-6 receptor signalling activity, which may produce more consistent associations in forward MR analyses.

### 4.5 Asymmetry in bidirectional causal effects

Notably, there was little overlap between traits identified in the forward and reverse MR analyses for IL-6. This asymmetry may reflect the tightly regulated nature of IL-6 production, for which few strong upstream genetic determinants exist, contrasted with the broader downstream metabolic consequences of IL-6 signalling, which are more robustly captured by genetic variation at the IL6R locus. Collectively, these findings suggest that while relatively few traits causally influence circulating IL-6 levels, genetically proxied IL-6 signalling may exert substantial downstream effects on lipid and lipoprotein metabolism, although many of these associations should be interpreted cautiously due to the limited number of available genetic instruments.

The absence of bidirectional MR effects when restricting analyses to *cis*-acting CRP instruments suggests that the bidirectional signals observed using broader instrument sets, as using *cis* and trans variants produced evidence for associations, are unlikely to reflect direct causal effects of CRP itself. Instead, these associations are more consistent with horizontal pleiotropy or heritable confounding, whereby *trans*-acting CRP variants influence metabolic traits through pathways independent of circulating CRP [78]. For example, variants associated with adiposity may be identified in large CRP GWAS due to shared genetic architecture, leading to apparent bidirectional associations between CRP and metabolic traits. Similar mechanisms have been proposed in previous genetic and MR studies [25,73].

Bidirectional MR provides additional evidence regarding the likely direction of causal relationships by testing associations in both directions. However, the temporal ordering and mechanistic relationships underlying these associations cannot always be fully resolved. Systemic inflammation is a highly interconnected and dynamic biological network. While circulating CRP and IL-6 are highly informative of inflammation, they only partially represent a broader system. Consequently, although MR can provide evidence consistent with causal effects on these biomarkers, it cannot fully resolve the complex relationships, feedback mechanisms, and temporal ordering of processes operating across the wider inflammatory network.

### 4.6 Strengths and limitations

By drawing on large, well-powered and widely-utilised GWAS available for CRP, IL-6, and a wide range of metabolomic, proteomic, and cardiometabolic traits, this study sought to maximise statistical power and improve the precision of effect estimates. Multiple complementary sensitivity analyses - including the weighted median, MR-Egger, and Steiger filtering -helped mitigate bias from horizontal pleiotropy and reinforced confidence in the inferred direction of causality. In addition, bidirectional MR provided further protection against reverse causation by allowing us to explicitly test causal relationships in both directions.

However, several limitations should be considered. Like all MR studies, our analyses rely on the validity of the instrumental variable assumptions, particularly the exclusion restriction assumption, whereby genetic variants influence the outcome only through the exposure of interest [56]. This assumption is particularly relevant in the present analyses because many investigated traits are biologically correlated and genetically pleiotropic, increasing the potential for horizontal pleiotropy and biased causal estimates.

Although horizontal pleiotropy cannot be completely excluded, several design choices were implemented to minimise this risk. For proteomic traits, primary analyses were restricted to *cis*-pQTLs, which are located within or near the gene encoding the corresponding protein [54,55]. These variants are generally considered less susceptible to horizontal pleiotropy than trans-pQTLs because they are more likely to influence protein abundance through regulation of the encoding gene [27–29]. Consequently, the cis-focused approach was intended to increase confidence that observed associations reflected effects mediated through the protein of interest rather than alternative biological pathways [53]. However, not all proteins had suitable cis-acting instruments, and many complex traits and diseases lack a clearly defined causal gene region from which cis instruments can be derived. As a result, many variants were excluded from primary analyses, reducing the number of available instruments and potentially limiting statistical power.

For all exposure types, genetic instruments were selected using the conventional genome-wide significance threshold (P < 5 × 10⁻⁸), which is widely used in MR studies to ensure robust instrument-exposure associations and minimise weak instrument bias [17]. Nevertheless, genome-wide significant variants may still violate instrumental variable assumptions through horizontal pleiotropy. To further mitigate this risk, we applied strict LD clumping criteria (R² < 0.001) to ensure independence of genetic instruments [20,79], Steiger filtering to remove variants more strongly associated with the outcome than the exposure [20], and multiple complementary sensitivity analyses, including weighted median and MR-Egger methods. Despite these precautions, residual pleiotropy and other sources of bias cannot be entirely excluded.

Additional measures were also implemented to reduce potential bias. Genetic instruments were selected using the conventional genome-wide significance threshold (P < 5 × 10⁻⁸), strict LD clumping criteria were applied (R² < 0.001) to ensure instrument independence [20,79], and Steiger filtering was used to remove variants more strongly associated with the outcome than the exposure [20]. Furthermore, complementary sensitivity analyses, including weighted median and MR-Egger methods, were performed where possible. Nevertheless, residual pleiotropy and other sources of bias cannot be entirely excluded.

Our findings are inherently influenced by the scope and quality of the GWAS data available. Analyses could only include traits with existing large scale summary statistics, the extent and interpretation of our results are shaped by the traits which have been comprehensively studied to date, as such autism ( N = 46350) and anxiety (N = 21761) were represented by relatively small GWAS sample sizes (although, these were the biggest available at time of analysis). For possibly underpowered GWAS, the combination of numerous tests and stringent correction for multiple comparisons increases the likelihood of false negatives, and true causal effects may therefore have gone undetected. As a result, areas with limited or less characterised GWAS may be underrepresented in the causal landscape we describe. In a similar manner, as our analyses are limited to the traits for which GWAS data exist, our conclusions are inherently shaped by the current coverage of metabolomic and proteomic platforms rather than the full biological landscape. The analyses were also restricted to European populations ancestry, reflecting the composition of the underlying GWAS datasets. Consequently, the generalisability of these findings to other ancestral groups may be limited. Future work could extend these analyses to large non-European biobanks, such as the Biobank Japan, China Kadoorie Biobank, and Million Veteran Program, to evaluate the consistency of the strongest associations across diverse populations.

Although MR can provide evidence for causal effects, the univariable analyses presented estimate total effects and do not investigate the temporal ordering or mechanistic position of these traits within broader inflammatory pathways. Future studies using multivariable MR, mediation analyses and network-based approaches may help disentangle these relationships.

Despite these limitations, the systematic design, use of large-scale genomic resources, and robust MR workflow collectively support the reliability of our findings and provide a strong foundation for future mechanistic and translational work.

### 4.7 Conclusion

This study provides a comprehensive overview of traits that may causally influence circulating IL-6 and CRP levels, helping to identify candidate upstream determinants of systemic inflammation while demonstrating that IL-6 and CRP are influenced by distinct metabolic and biological processes. While adiposity is as a shared causal driver of both biomarkers, CRP was influenced by a broad spectrum of metabolic, lipidomic, amino acid, and proteomic traits, consistent with its role as an integrated downstream readout of systemic inflammatory burden [1]. In contrast, IL-6 levels are shaped by a more constrained upstream profile, driven primarily by adiposity and HDL-related pathways, reflecting its tighter regulatory control and more context-dependent signalling biology [48]. These findings highlight that CRP and IL-6 are not interchangeable indicators of systemic inflammation but instead occupy overlapping yet mechanistically distinct positions within the inflammatory-metabolic network, with differing implications for causal inference and intervention.

By directly contrasting upstream and downstream determinants of IL-6 and CRP within a bidirectional framework, this study moves beyond isolated MR analyses and highlights the importance of disentangling causes from consequences of inflammation. Future studies should aim to refine these pathways, disentangle signalling-specific mechanisms, particularly so for IL-6, and assess whether interventions (pharmaceutical or lifestyle) targeting upstream metabolic traits such as adiposity and triglyceride rich lipoproteins could be used to meaningfully reduce systemic inflammation, the increase of which is in turn associated with multiple chronic disease outcomes.

## Data Availability

All data produced in the present work are contained in the manuscript and
All data produced are available online at https://doi.org/10.6084/m9.figshare.34420311

https://github.com/shepherdfreya/DeterminingSystemicInflammation

## Data availability

## Source data

The GWAS summary statistics analysed in this study are publicly available from the sources cited in the text and listed in Extended data Tables 1 and 2 [22]. These tables provide dataset identifiers and access links.

## Extended data

Extended data for “Prioritising determinants of systemic inflammation across molecular, physiological and disease phenotypes” is available at Figshare [https://doi.org/10.6084/m9.figshare.34420311] [22].

This project contains the following extended data:

- Supplementary tables.
- Supplementary methods.
- STROBE-MR checklist.

The extended data are available under the terms of the Creative Commons Attribution 4.0 International (CC BY 4.0) licence.

## Software availability

Software available from: https://github.com/shepherdfreya/DeterminingSystemicInflammation

Source code available from: https://github.com/shepherdfreya/DeterminingSystemicInflammation

Archived source code at time of publication: https://doi.org/10.5281/zenodo.23416330 [80].

License: MIT License.

The repository contains the analytical code for this study. Details of the software and packages used, including version numbers, are provided in the extended data [22] and the GitHub repository.

## References

[1] Pepys MB, Hirschfield GM. C-reactive protein: a critical update. J Clin Invest 2003;111:1805–12. 10.1172/JCI18921.

[2] Alivernini S, Firestein GS, McInnes IB. The pathogenesis of rheumatoid arthritis. Immunity 2022;55:2255–70. 10.1016/j.immuni.2022.11.009.

[3] Cupido AJ, Asselbergs FW, Natarajan P, Ridker PM, Hovingh GK, Schmidt AF. Dissecting the IL-6 pathway in cardiometabolic disease: A Mendelian randomization study on both IL6 and IL6R. Br J Clin Pharmacol 2022;88:2875–84. 10.1111/bcp.15191.

[4] Khandaker GM, Pearson RM, Zammit S, Lewis G, Jones PB. Association of Serum Interleukin 6 and C-Reactive Protein in Childhood With Depression and Psychosis in Young Adult Life: A Population-Based Longitudinal Study. JAMA Psychiatry 2014;71:1121–8. 10.1001/jamapsychiatry.2014.1332.

[5] Lustgarten MS, Fielding RA. Metabolites Associated With Circulating Interleukin-6 in Older Adults. J Gerontol A Biol Sci Med Sci 2017;72:1277–83. 10.1093/gerona/glw039.

[6] D’Alessandro A, Thomas T, Dzieciatkowska M, Hill RC, Francis RO, Hudson KE, et al. Serum Proteomics in COVID-19 Patients: Altered Coagulation and Complement Status as a Function of IL-6 Level. J Proteome Res 2020;19:4417–27. 10.1021/acs.jproteome.0c00365.

[7] Hunter CA, Jones SA. IL-6 as a keystone cytokine in health and disease. Nat Immunol 2015;16:448–57. 10.1038/ni.3153.

[8] Tanaka T, Narazaki M, Kishimoto T. IL-6 in Inflammation, Immunity, and Disease. Cold Spring Harbor Perspectives in Biology 2014;6:a016295. 10.1101/cshperspect.a016295.

[9] Foley ÉM, Parkinson JT, Mitchell RE, Turner L, Khandaker GM. Peripheral blood cellular immunophenotype in depression: a systematic review and meta-analysis. Mol Psychiatry 2023;28:1004–19. 10.1038/s41380-022-01919-7.

[10] Singh B, Goyal A, Patel BC. C-Reactive Protein: Clinical Relevance and Interpretation. StatPearls, Treasure Island (FL): StatPearls Publishing; 2025.

[11] Henein MY, Vancheri S, Longo G, Vancheri F. The Role of Inflammation in Cardiovascular Disease. Int J Mol Sci 2022;23:12906. 10.3390/ijms232112906.

[12] Devaraj S, Singh U, Jialal I. Human C-reactive protein and the metabolic syndrome. Curr Opin Lipidol 2009;20:182–9. 10.1097/MOL.0b013e32832ac03e.

[13] Ni P, Yu M, Zhang R, Cheng C, He M, Wang H, et al. Dose-response association between C-reactive protein and risk of all-cause and cause-specific mortality: a systematic review and meta-analysis of cohort studies. Annals of Epidemiology 2020;51:20–27.e11. 10.1016/j.annepidem.2020.07.005.

[14] Chamberlain SR, Cavanagh J, de Boer P, Mondelli V, Jones DNC, Drevets WC, et al. Treatment-resistant depression and peripheral C-reactive protein. Br J Psychiatry 2019;214:11–9. 10.1192/bjp.2018.66.

[15] Pitharouli MC, Hagenaars SP, Glanville KP, Coleman JRI, Hotopf M, Lewis CM, et al. Elevated C-Reactive Protein in Patients With Depression, Independent of Genetic, Health, and Psychosocial Factors: Results From the UK Biobank. American Journal of Psychiatry 2021;178:522–9. 10.1176/appi.ajp.2020.20060947.

[16] Davey Smith G, Ebrahim S. ‘Mendelian randomization’: can genetic epidemiology contribute to understanding environmental determinants of disease?*. International Journal of Epidemiology 2003;32:1–22. 10.1093/ije/dyg070.

[17] Sanderson E, Glymour MM, Holmes MV, Kang H, Morrison J, Munafò MR, et al. Mendelian randomization. Nat Rev Methods Primers 2022;2:6. 10.1038/s43586-021-00092-5.

[18] Davey Smith G, Hemani G. Mendelian randomization: genetic anchors for causal inference in epidemiological studies. Human Molecular Genetics 2014;23:R89–98. 10.1093/hmg/ddu328.

[19] Bowden J, Davey Smith G, Burgess S. Mendelian randomization with invalid instruments: effect estimation and bias detection through Egger regression. International Journal of Epidemiology 2015;44:512–25. 10.1093/ije/dyv080.

[20] Burgess S, Davey Smith G, Davies NM, Dudbridge F, Gill D, Glymour MM, et al. Guidelines for performing Mendelian randomization investigations: update for summer 2023. Wellcome Open Res 2019;4:186. 10.12688/wellcomeopenres.15555.3.

[21] Howe LJ, Tudball M, Davey Smith G, Davies NM. Interpreting Mendelian-randomization estimates of the effects of categorical exposures such as disease status and educational attainment. Int J Epidemiol 2022;51:948–57. 10.1093/ije/dyab208.

[22] Prioritising determinants of systemic inflammation across molecular, physiological and disease phenotypes. Figshare 2026. 10.6084/m9.figshare.34420311.v1.

[23] Shepherd F, Slaney C, Jones HJ, Dardani C, Stergiakouli E, Sanderson ECM, et al. Prioritising determinants of systemic inflammation across molecular, physiological and disease phenotypes. 2026. 10.64898/2026.04.10.26350510.

[24] Ahluwalia TS, Prins BP, Abdollahi M, Armstrong NJ, Aslibekyan S, Bain L, et al. Genome-wide association study of circulating interleukin 6 levels identifies novel loci. Hum Mol Genet 2021;30:393–409. 10.1093/hmg/ddab023.

[25] Ligthart S, Vaez A, Võsa U, Stathopoulou MG, de Vries PS, Prins BP, et al. Genome Analyses of >200,000 Individuals Identify 58 Loci for Chronic Inflammation and Highlight Pathways that Link Inflammation and Complex Disorders. Am J Hum Genet 2018;103:691–706. 10.1016/j.ajhg.2018.09.009.

[26] Said S, Pazoki R, Karhunen V, Võsa U, Ligthart S, Bodinier B, et al. Genetic analysis of over half a million people characterises C-reactive protein loci. Nat Commun 2022;13:2198. 10.1038/s41467-022-29650-5.

[27] Tambets R, Jesse M, Kronberg J, van der Graaf A, Abner E, Võsa U, et al. Genetic analysis of circulating metabolic traits in 619,372 individuals. Nature 2026:1–8. 10.1038/s41586-026-10532-5.

[28] Sun BB, Chiou J, Traylor M, Benner C, Hsu Y-H, Richardson TG, et al. Plasma proteomic associations with genetics and health in the UK Biobank. Nature 2023;622:329–38. 10.1038/s41586-023-06592-6.

[29] Yengo L, Sidorenko J, Kemper KE, Zheng Z, Wood AR, Weedon MN, et al. Meta-analysis of genome-wide association studies for height and body mass index in ∼700000 individuals of European ancestry. Hum Mol Genet 2018;27:3641–9. 10.1093/hmg/ddy271.

[30] Graham SE, Clarke SL, Wu K-HH, Kanoni S, Zajac GJM, Ramdas S, et al. The power of genetic diversity in genome-wide association studies of lipids. Nature 2021;600:675–9. 10.1038/s41586-021-04064-3.

[31] Chen J, Spracklen CN, Marenne G, Varshney A, Corbin LJ, Luan J, et al. The trans-ancestral genomic architecture of glycemic traits. Nat Genet 2021;53:840–60. 10.1038/s41588-021-00852-9.

[32] Suzuki K, Hatzikotoulas K, Southam L, Taylor HJ, Yin X, Lorenz KM, et al. Genetic drivers of heterogeneity in type 2 diabetes pathophysiology. Nature 2024;627:347–57. 10.1038/s41586-024-07019-6.

[33] Keaton JM, Kamali Z, Xie T, Vaez A, Williams A, Goleva SB, et al. Genome-wide analysis in over 1 million individuals of European ancestry yields improved polygenic risk scores for blood pressure traits. Nat Genet 2024;56:778–91. 10.1038/s41588-024-01714-w.

[34] Yaghootkar H, Zhang Y, Spracklen CN, Karaderi T, Huang LO, Bradfield J, et al. Genetic Studies of Leptin Concentrations Implicate Leptin in the Regulation of Early Adiposity. Diabetes 2020;69:2806–18. 10.2337/db20-0070.

[35] Backman JD, Li AH, Marcketta A, Sun D, Mbatchou J, Kessler MD, et al. Exome sequencing and analysis of 454,787 UK Biobank participants. Nature 2021;599:628–34. 10.1038/s41586-021-04103-z.

[36] Verma A, Huffman JE, Rodriguez A, Conery M, Liu M, Ho Y-L, et al. Diversity and scale: Genetic architecture of 2068 traits in the VA Million Veteran Program. Science 2024;385:eadj1182. 10.1126/science.adj1182.

[37] Pedersen EM, Agerbo E, Plana-Ripoll O, Steinbach J, Krebs MD, Hougaard DM, et al. ADuLT: An efficient and robust time-to-event GWAS. Nat Commun 2023;14:5553. 10.1038/s41467-023-41210-z.

[38] Demontis D, Walters GB, Athanasiadis G, Walters R, Therrien K, Nielsen TT, et al. Genome-wide analyses of ADHD identify 27 risk loci, refine the genetic architecture and implicate several cognitive domains. Nat Genet 2023;55:198–208. 10.1038/s41588-022-01285-8.

[39] Otowa T, Hek K, Lee M, Byrne EM, Mirza SS, Nivard MG, et al. Meta-analysis of genome-wide association studies of anxiety disorders. Mol Psychiatry 2016;21:1391–9. 10.1038/mp.2015.197.

[40] Als TD, Kurki MI, Grove J, Voloudakis G, Therrien K, Tasanko E, et al. Depression pathophysiology, risk prediction of recurrence and comorbid psychiatric disorders using genome-wide analyses. Nat Med 2023;29:1832–44. 10.1038/s41591-023-02352-1.

[41] O’Connell KS, Koromina M, van der Veen T, Boltz T, David FS, Yang JMK, et al. Genomics yields biological and phenotypic insights into bipolar disorder - Nature. Nature 2025;639:968–75. 10.1038/s41586-024-08468-9.

[42] Trubetskoy V, Pardiñas AF, Qi T, Panagiotaropoulou G, Awasthi S, Bigdeli TB, et al. Mapping genomic loci implicates genes and synaptic biology in schizophrenia. Nature 2022;604:502–8. 10.1038/s41586-022-04434-5.

[43] Jansen IE, Savage JE, Watanabe K, Bryois J, Williams DM, Steinberg S, et al. Genome-wide meta-analysis identifies new loci and functional pathways influencing Alzheimer’s disease risk - Nature Genetics. Nature Genetics 2019;51:404–13. 10.1038/s41588-018-0311-9.

[44] Newbury A, Elhussein A, Gürsoy G. Multi-domain rule-based phenotyping algorithms enable improved GWAS signal. Npj Digit Med 2025;8:499. 10.1038/s41746-025-01815-8.

[45] Crouch DJM, Inshaw JRJ, Robertson CC, Ng E, Zhang J, Chen W, et al. Bayesian Effect Size Ranking to Prioritise Genetic Risk Variants in Common Diseases for Follow-Up Studies. Genetic Epidemiology 2025;49:e22608. 10.1002/gepi.22608.

[46] Loya H, Kalantzis G, Cooper F, Palamara PF. A scalable variational inference approach for increased mixed-model association power. Nat Genet 2025;57:461–8. 10.1038/s41588-024-02044-7.

[47] Shigesi N, Harris HR, Fang H, Ndungu A, Lincoln MR, The International Endometriosis Genome Consortium, et al. The phenotypic and genetic association between endometriosis and immunological diseases. Hum Reprod 2025;40:1195–209. 10.1093/humrep/deaf062.

[48] Hamilton FW, Thomas M, Arnold D, Palmer T, Moran E, Mentzer AJ, et al. Therapeutic potential of IL6R blockade for the treatment of sepsis and sepsis-related death: A Mendelian randomisation study. PLoS Med 2023;20:e1004174. 10.1371/journal.pmed.1004174.

[49] The COVID-19 Host Genetics Initiative. The COVID-19 Host Genetics Initiative, a global initiative to elucidate the role of host genetic factors in susceptibility and severity of the SARS-CoV-2 virus pandemic. Eur J Hum Genet 2020;28:715–8. 10.1038/s41431-020-0636-6.

[50] Xiang R, Ben-Eghan C, Liu Y, Roberts D, Ritchie S, Lambert SA, et al. Genome-wide analyses of variance in blood cell phenotypes provide new insights into complex trait biology and prediction - Nature Communications. Nature Communications 2025;16:4260. 10.1038/s41467-025-59525-4.

[51] Purcell S, Neale B, Todd-Brown K, Thomas L, Ferreira MAR, Bender D, et al. PLINK: A Tool Set for Whole-Genome Association and Population-Based Linkage Analyses. Am J Hum Genet 2007;81:559–75. 10.1086/519795.

[52] 1000 Genomes Project Consortium, Auton A, Brooks LD, Durbin RM, Garrison EP, Kang HM, et al. A global reference for human genetic variation. Nature 2015;526:68–74. 10.1038/nature15393.

[53] Holmes MV, Richardson TG, Ference BA, Davies NM, Davey Smith G. Integrating genomics with biomarkers and therapeutic targets to invigorate cardiovascular drug development. Nat Rev Cardiol 2021;18:435–53. 10.1038/s41569-020-00493-1.

[54] Swerdlow DI, Kuchenbaecker KB, Shah S, Sofat R, Holmes MV, White J, et al. Selecting instruments for Mendelian randomization in the wake of genome-wide association studies. Int J Epidemiol 2016;45:1600–16. 10.1093/ije/dyw088.

[55] Dardani C, Robinson JW, Jones HJ, Rai D, Stergiakouli E, Grove J, et al. Immunological drivers and potential novel drug targets for major psychiatric, neurodevelopmental, and neurodegenerative conditions. Mol Psychiatry 2025:1–10. 10.1038/s41380-025-03032-x.

[56] Hemani G, Tilling K, Smith GD. Orienting the causal relationship between imprecisely measured traits using GWAS summary data. PLOS Genetics 2017;13:e1007081. 10.1371/journal.pgen.1007081.

[57] Bowden J, Smith GD, Haycock PC, Burgess S. Consistent Estimation in Mendelian Randomization with Some Invalid Instruments Using a Weighted Median Estimator. Genetic Epidemiology 2016;40:304. 10.1002/gepi.21965.

[58] Stock JH, Yogo M. Testing for Weak Instruments in Linear IV Regression 2004.

[59] Slaney C, Sallis HM, Jones HJ, Dardani C, Tilling K, Munafò MR, et al. Association between inflammation and cognition: Triangulation of evidence using a population-based cohort and Mendelian randomization analyses. Brain, Behavior, and Immunity 2023;110:30–42. 10.1016/j.bbi.2023.02.010.

[60] Zhang L, Omarov M, Xu L, deGoma E, Natarajan P, Georgakis MK. IL6 genetic perturbation mimicking IL-6 inhibition is associated with lower cardiometabolic risk. Nat Cardiovasc Res 2025;4:1172–86. 10.1038/s44161-025-00700-7.

[61] The Interleukin-6 Receptor Mendelian Randomisation Analysis (IL6R MR) Consortium. The interleukin-6 receptor as a target for prevention of coronary heart disease: a mendelian randomisation analysis. The Lancet 2012;379:1214–24. 10.1016/S0140-6736(12)60110-X.

[62] E S, G DS, F W, J B. An examination of multivariable Mendelian randomization in the single-sample and two-sample summary data settings. International Journal of Epidemiology 2019;48. 10.1093/ije/dyy262.

[63] Carter AR, Sanderson E, Hammerton G, Richmond RC, Davey Smith G, Heron J, et al. Mendelian randomisation for mediation analysis: current methods and challenges for implementation. Eur J Epidemiol 2021;36:465–78. 10.1007/s10654-021-00757-1.

[64] Hemani G, Zheng J, Elsworth B, Wade KH, Haberland V, Baird D, et al. The MR-Base platform supports systematic causal inference across the human phenome. eLife 2018;7:e34408. 10.7554/eLife.34408.

[65] Hemani G, Elsworth B, Palmer T, Rasteiro R. ieugwasr: Interface to the OpenGWAS Database API. 2025. 10.32614/CRAN.package.ieugwasr.

[66] Skrivankova VW, Richmond RC, Woolf BAR, Davies NM, Swanson SA, VanderWeele TJ, et al. Strengthening the reporting of observational studies in epidemiology using mendelian randomisation (STROBE-MR): explanation and elaboration. BMJ 2021;375:n2233. 10.1136/bmj.n2233.

[67] Ferreira RC, Freitag DF, Cutler AJ, Howson JMM, Rainbow DB, Smyth DJ, et al. Functional IL6R 358Ala Allele Impairs Classical IL-6 Receptor Signaling and Influences Risk of Diverse Inflammatory Diseases. PLOS Genetics 2013;9:e1003444. 10.1371/journal.pgen.1003444.

[68] Coppack SW. Pro-inflammatory cytokines and adipose tissue. Proceedings of the Nutrition Society 2001;60:349–56. 10.1079/PNS2001110.

[69] Timpson NJ, Nordestgaard BG, Harbord RM, Zacho J, Frayling TM, Tybjærg-Hansen A, et al. C-reactive protein levels and body mass index: Elucidating direction of causation through reciprocal Mendelian randomization. Int J Obes (Lond) 2011;35:300–8. 10.1038/ijo.2010.137.

[70] Rose-John S. Interleukin-6 signalling in health and disease. F1000Res 2020;9:F1000 Faculty Rev-1013. 10.12688/f1000research.26058.1.

[71] Calabrese LH, Rose-John S. IL-6 biology: implications for clinical targeting in rheumatic disease. Nat Rev Rheumatol 2014;10:720–7. 10.1038/nrrheum.2014.127.

[72] Garbers C, Monhasery N, Aparicio-Siegmund S, Lokau J, Baran P, Nowell MA, et al. The interleukin-6 receptor Asp358Ala single nucleotide polymorphism rs2228145 confers increased proteolytic conversion rates by ADAM proteases. Biochimica et Biophysica Acta (BBA) - Molecular Basis of Disease 2014;1842:1485–94. 10.1016/j.bbadis.2014.05.018.

[73] Sanderson E, Rosoff D, Palmer T, Tilling K, Smith GD, Hemani G. Heritable confounding in Mendelian randomization studies: structure, consequences and relevance for gene-environment equivalence 2024. 10.1101/2024.09.05.24312293.

[74] Rader DJ, Hovingh GK. HDL and cardiovascular disease. The Lancet 2014;384:618–25. 10.1016/S0140-6736(14)61217-4.

[75] Ridker PM, Rifai N, Rose L, Buring JE, Cook NR. Comparison of C-reactive protein and low-density lipoprotein cholesterol levels in the prediction of first cardiovascular events. N Engl J Med 2002;347:1557–65. 10.1056/NEJMoa021993.

[76] G HB, Rao VS, Kakkar VV. Friend Turns Foe: Transformation of Anti-Inflammatory HDL to Proinflammatory HDL during Acute-Phase Response. Cholesterol 2011;2011:274629. 10.1155/2011/274629.

[77] Ouyang FW, Chiang H-H, Hsu W-L, Tsai M-H, Huang C-Y, Remaley AT, et al. Dysfunctional high-density lipoprotein: an updated review. Front Cardiovasc Med n.d.;12:1713387. 10.3389/fcvm.2025.1713387.

[78] Burgess S, Thompson SG. Interpreting findings from Mendelian randomization using the MR-Egger method. Eur J Epidemiol 2017;32:377–89. 10.1007/s10654-017-0255-x.

[79] Burgess S, Thompson SG. Mendelian Randomization. 0 ed. Chapman and Hall/CRC; 2015. 10.1201/b18084.

